# Using a self-attention architecture to automate valence categorization of French teenagers’ free descriptions of their family relationships. A proof of concept

**DOI:** 10.1101/2023.01.16.23284557

**Authors:** Mohammed Sedki, Nathan Vidal, Paul Roux, Caroline Barry, Mario Speranza, Bruno Falissard, Eric Brunet-Gouet

## Abstract

This paper proposes a proof of concept of using natural language processing techniques to categorize valence of family relationships described in free texts written by french teenagers. The proposed study traces the evolution of techniques for word embedding. After decomposing the different texts in our possession into short texts composed of sentences and manual labeling, we tested different word embedding scenarios to train a multi-label classification model where a text can take several labels : labels describing the family link between the teenager and the person mentioned in the text and labels describing the teenager’s relationship with them positive/negative/neutral valence). The natural baseline for word vector representation of our texts is to build a TF-IDF and train classical classifiers (Elasticnet logistic regression, gradient boosting, random forest, support vector classifier) after selecting a model by cross validation in each class of machine learning models. We then studied the strengths of word-vectors embeddings by an advanced language representation technique via the CamemBERT transformer model, and, again, used them with classical classifiers to compare their respective performances. The last scenario consisted in augmenting the CamemBERT with output dense layers (perceptron) representing a classifier adapted to the multi-label classification and fine-tuning the CamemBERT original layers. The optimal fine-tuning depth that achieves a bias-variance trade-off was obtained by a cross-validation procedure. The results of the comparison of the three scenarios on a test dataset show a clear improvement of the classification performances of the scenario with fine-tuning beyond the baseline and of a simple vectorization using CamemBERT without fine-tuning. Despite the moderate size of the dataset and the input texts, fine-tuning to an optimal depth remains the best solution to build a classifier.

## 1 Introduction

Research on family interactions and their perception by adolescents is a major issue for the management of patients in child psychiatry. Complex epistemological and methodological questions are raised by the numerous works in the concerned disciplines, for which it seems interesting to bring new tools from artificial intelligence technologies, in particular from automated language processing. Indeed, the considerable increase in the interaction of teenagers with digital tools can be investigated with methods of analysis of verbal or textual data. In the following we confront family research on teenagers populations, with the state-of-the-art natural language processing (NLP) and “sentiment analysis” (i.e. determining the emotional and subjective valence from a text) and try to produce new tools with the aim of fostering larger scale protocols. Let’s note that research based on narratives is scarce and represents only 2% of the available literature in NLP applications to research on mental health conditions detection, the vast majority being based on social media database [1].

Despite the wealth of literature in family research, no consensus has been established on the variables and constructs to describe quantitatively family relationships [2]. The most relevant theoretical frameworks focus on family histories (family development theory), systemic relationships between family members, family relationships with the environment, attachment relationships, social learning by children, etc. These different approaches are reflected in a wide variety of experimental methods, but the depth of the epistemological differences constitutes a serious pitfall for the comparison of research results. In order to overcome the differences, Falissard and colleagues have developed a common tool between sociologists, psychoanalysts and adolescent psychiatrists to be applied to the free discourse that adolescents may hold about their own family relationships [2]. 194 French adolescents (mean age : 14.7(*s.d*. = 2) years, 51% girls) were recruited to produce a corpus of descriptions of their family relationships (mean text length : 232(*s.d*. = 129) words). The instructions were : “*In the next half hour, would you please write as freely as you wish about your relationships in your family, explaining how things are. All that you write is anonymous and no parent or person from your school will read it* “. These short texts were analysed and rated by blind raters across 18 dimensions (affective environment, conflict, injustice, support, positive/negative relations, etc.) as decided by an expert consensus. After a careful metrological investigation on the items, an exploratory factorial analysis was conducted and resulted in a unifactorial solution accounting for more than half the variance. This solution emphasized the positive/negative valence of relationships with other family members. Thus, it appears through this interdisciplinary research that relational valence constitutes a key element of the family descriptions produced by adolescents and thus undoubtedly of their mental representations of them. [2] also argue in favor of using this dimension as a primary endpoint in future interventional research. We add to their conclusion, that if it is accepted that the valence of relationships between individuals cannot be ignored in any family assessment, this provides a strong rationale for sentiment analysis studies (see [3] for an overview of this NLP application domain) aimed at automating the analysis of adolescent free speech or writings. To exemplify the possibility to use NLP to classify texts from first-episodes patients with schizophrenia compared with healthy controls, Gutierrez et al. combined methaphoricity assessment and sentiment analysis (emotional valence of texts) to train a Recursive Neural Network classifier [4].

### Supervised learning to analyze free texts’ contents

Dealing with labelled textual data in a supervised learning problem raises the challenge to find an convenient way to represent texts contents so that prediction algorithms could process input datasets. Extracting features from the text that will feed the classifiers is the starting point for using a supervised learning model. A first solution has been proposed for the preparation and digitization of a text as a numerical vector called Term Frequency (TF), corresponding to the count of the occurrences of each term within the text, and its variant TF-Inverse-Document-Frequency (TF-IDF), in which counts of occurrences are inversely weighted by their frequencies in the corpus [5, 6]. This technique has been popularized by the work of [7] in unsupervised document classification. TF and TF-IDF can be seen as simple text embedding methods to feed classical supervised learning models ranging from penalized logistic regression to tree models like gradient boosting and random forest. Word embedding of textual data using TF or TF-IDF does not allow for the consideration of ordered word sequences in a text and is invariant to permutation of words. Taking into account the words order in a text is a real challenge for improving the predictive performance of supervised learning models.

Deep-learning approaches brought new efficient way to achieve supervised learning NLP tasks using a general back-propagation of error mechanism, and benefiting from large corpus of training data, and is now commonly investigated in emotion labeling tasks [8]. Successive improvements, allowing for a better handling of word-order, of long-term dependency as well of gradient vanishing problems, used Recurrent Neural Networks (RNN), Long Short-Term Memory (LSTM), and Self-attention or Transformers architectures. As noted by [9], deep learning methods were only recently introduced in NLP literature applied to mental health. Some authors advocated for the use of fine coherence and syntactic NLP processing to classify diagnosis such as psychosis [10].

Recently a solution showing a qualitative leap in terms of performance for embedding textual data was proposed by Google AI via the implementation of a sophisticated neural architecture called BERT [11](*Bidirectional Encoder Representations from Transformers*). This model implements an attentional mechanism that consists in weighting the input vectors in order to form a context vector (i.e. the 12 self-attention heads acting in parallel) used to process each word through feed-forward network, layer normalization and dropout mechanisms.

A work based on self-attentional architectures described the use of such NLP models to classify 209000 texts from the GermEval 2020 task[12]. Pieces of free texts describing a person situation, feelings and actions from simple drawings of the Operant Motive Test [13] were classified by trained psychologists into five possible motives and rated into six possible levels. Several architectures were compared such as supervised autoencoders, fully connected neural networks, and transformers (BERT, XLM, DistilBERT), with respect to a baseline consisting of a Support Vector Classifier of TF-IDF text representation.

Interestingly, the best classifier performance, i.e. F1-scores of 0.69, was found with a simple BERT model. Another work showed that BERT model could be successfully used to classify social media sentences into five basic emotions, with a high macro F1-score of 0.83 [14].

One question raised by the use of self-attentive models concerns the lexical, syntactic and semantic features processed by the different processing layers (the BERT model is composed of 11 layers of identical structure, themselves composed of several sub-layers). Since the encoder has the duty to transform N word-vectors of 768 dimensions into a single output vector of the same size, we hypothesize that each successive layer progressively reduces dimensionality while increasing in abstraction. Understanding deep-learning models is generally complicated and is a research question in itself, far beyond the scope of this work. As discussed by Jain et al., one would think that attentional weights are directly related to the importance given to inputs, which would help the interpretability of these models, but experimentation shows that this is not the case [15]. Similarly, a close examination of BERT’s attentional weights in the aforementioned GermEval classification task shows that the transformer pays more attention to form features (i.e. use of personal pronouns, stop words, negation, punctuation marks, unknown words, and some conjugation styles) than to content words [12]. While interpretability of attentional weights proves difficult, other authors have conducted a layer by layer examination of the structure of transformers by probing the corresponding hidden outputs. In question answering tasks, it was shown that successive layers support processing allowing for named entity extraction, coreference resolution, relation classification, and supporting fact extraction [16]. Although our use of transformer differs with respect to the task, the question is of importance in order to select if and how deep fine-tuning of pretrained models should be applied to models to achieve tasks akin to sentiment analysis.

## 2 Objectives

From this brief overview, we draw several objectives in the present study. We will challenge as a proof-of-concept the use of state-of-the-art NLP learning techniques on the corpus of adolescent texts described in Falissard et al. [2]. As described above, this corpus exhibits maximal variance along the positive vs. negative relationships dimension. In order to perform an analysis of their valence, we will transform text segments from the corpus with a *BERT* -derived model [11], pre-trained on a French database [17], into vector on which classification techniques can be applied. More precisely, we will be interested in classifying the valence, like in sentiment analysis, but also the categories of the people described in the text segments (i.e. mother, father, sister, the respondent him/herself, etc.). We will compare the classification performances of the classical algorithms : elastic net logistic regression, gradient boosting classifier, random forest, support vector classifier. In addition, we will test the added value of transformers’ embedding with a text vectorization method based on TF-IDF that does not take into account word order information. To go further in our understanding of machine learning usability in our field, we will test the interest of fine-tuning the upper layers of the transformer. We tackle the question of the categories of semantic information (i.e. person categories and/or relational valence) the transformer actually encodes in order to determine whether this information is represented in the output of the transformer, and usable for prediction. In addition, we raise the question of the level of fine-tuning that could improve classification performance.

## 3 Methods

In this section, we describe the different steps of data preparation, classification and finally comparison of the methods. The labeled textual datasets will be common to all evaluations. We will then transform the texts into vectors either by the TF-IDF method or by applying an attentional model. Then, several families of classical classification models are used, each one having been adjusted for its main hyperparameters. We also use the possibility to extend the attentional model by a perceptron in order to obtain a prediction. The attentional model itself will be compared using different depths of fine-tuning. Figures 1 and 2 illustrate the different computations used in the experiments.

**Figure 1.**
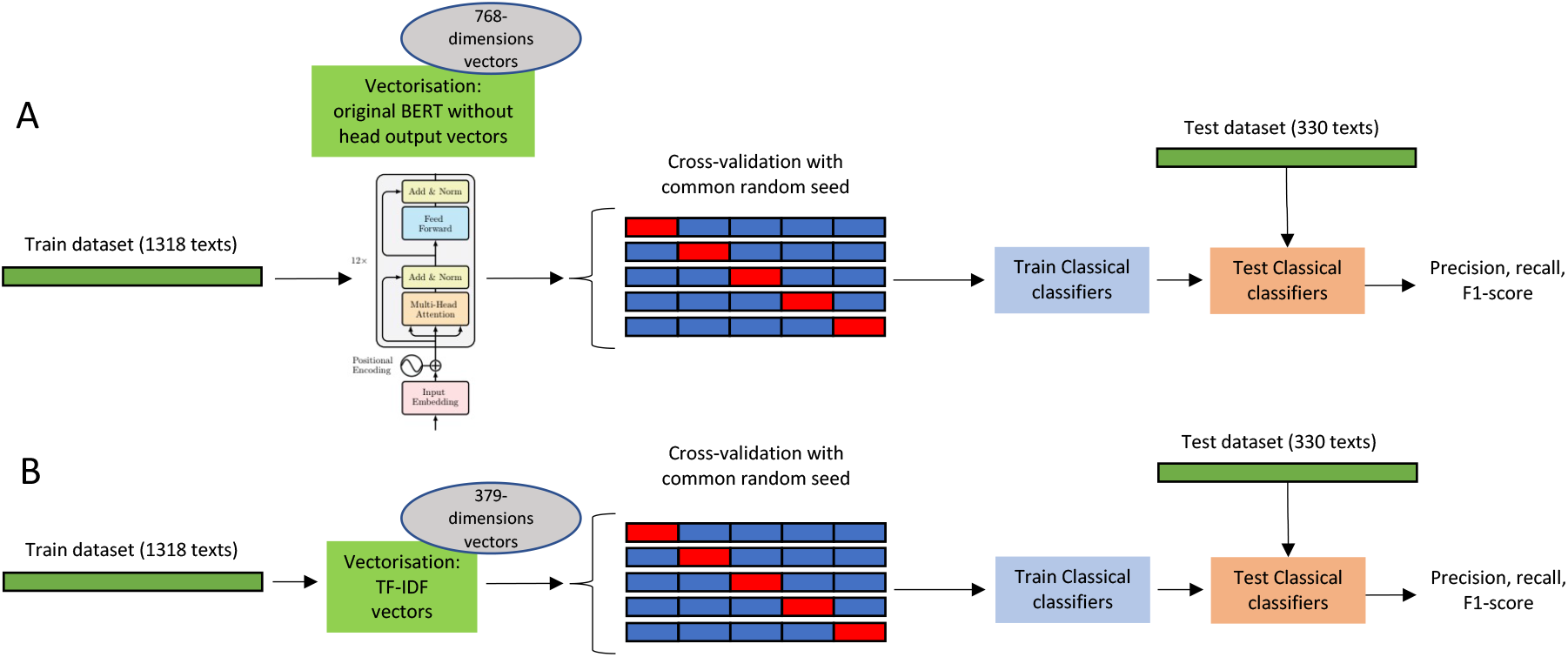
Design of the experiments. (A) Classical prediction algorithms (Elasticnet logistic regression, gradient boosting, random forest, support vector classifier) were evaluated using a 5-folds cross-validation procedure on the training dataset, with a common random seed to ensure that training and testing dataset are the same from an experiment to another. These classifiers are fed with CamemBERT’s 768-dimensions vectors. (B) TF-IDF vectors are used to evaluate the classical prediction algorithms with the same cross-validation procedure based on the same random seed.

**Figure 2.**
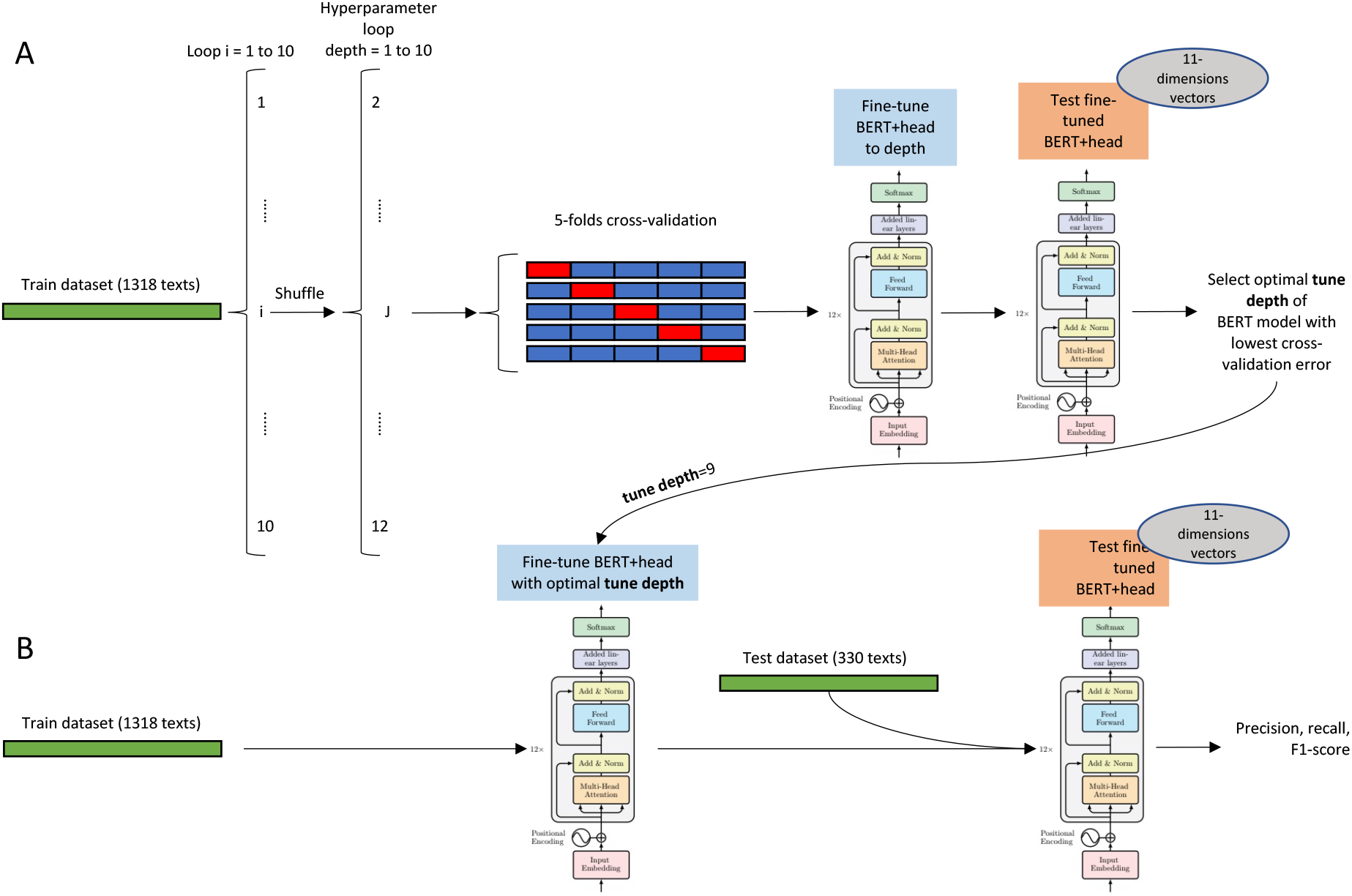
Design of the experiments. (A) To determine the optimal `tune depth` to fine-tune BERT and the perceptron on-top, a 5-folds cross-validation procedure, repeated ten times, was used to train the model and then measure prediction error. In subsequent cross-validation computations, this hyper-parameter was used to train CamemBERT. (B) Assessment of fine-tuned BERT was achieved on the training dataset with previously fixed learning hyper-parameters. Precision, recall and F1-Scores are obtained.

### 3.1 Data preparation and labelling

Texts from 194 teenagers from [2] were used to generate a set of 1648 text segments (8.4 segments in average per text ; max segment length : 345 chars. Please refer to this article for description of data collection and related legal agreements from the IRB. Note that we will use the term *segment* in the following to refer to these pieces of text). Each one was composed of one or several sentences depending on the presence of referential pronouns that have to be taken into account as a context to understand an assertion. For instance, “I have good relationships with my mother and my father with whom I live.” and “I live with my mother and father. I have good relationships with them.” are both taken as a single text segment to be labelled and to be processed. Thus, each segment could be unambiguously interpreted either by a human or an automated semantic analysis.

Once the segmentation was carried out, segments were labelled with 11 binary tags by one of the authors (EBG) according to simple criteria on the valence (*Valence*) and type of information given and concerning the people involved in the relationship (*Subject*) described. Whenever the segment contained information on relational valence, it was rated as positive (*+*), negative (*-*) or neutral (*0*). Positive relationships refer to a good understanding, an expression of positive affect, cooperation between the teenager and the subject (i.e. “I get along very well with my mother” or “My sister is like a close friend”). Negative relationships correspond to conflicts, disagreement, absence of a normal relationship, etc. (“My father is aggressive with me” or “My sister doesn’t talk to me, she’s a stranger to me”). Finally, the neutrality (*0*) of the statement is identified when the text implies an emotional relationship between the subject and another one (“I live with my mother and I see my father all the time”) and/or contains both positive and negative elements (ambivalent or ambiguous feelings) and does not allow for a clear valence to be inferred (“My father is nice to me but most of the time I can’t stand him”). In the absence of valence information, the text was considered as informative (*Info*) about the habits or living conditions of the persons when they did clearly imply a form of relationship (for instance, “My parents eat together in the evening with the children” do not imply that the respondent is involved and describe more a way of life than a relational involvement of the persons). The subjects described in a segment have been labeled as follows : the respondent (*Me*), “Mother”, “Father”, “Sister”, “Brother”, “Family member”, “Others”. As this labeling method was intended to be simple and coarse, this procedure did not require any linguistic expertise other than being proficient in the corresponding native language.

Finally, the dataset consisted of 1648 items with their 11 labels and was randomly split into a training dataset and a test dataset of sizes 1318 and 330, respectively. The same random generator seed was used to have the same test dataset for all the compared model families.

### 3.2 Text vectorization

Two text vectorization methods were applied, TF-IDF and Transformers, in order to feed the classical classifiers.

#### 3.2.1 Term-frequency-inverse-document

For any word w in a text segment t, TF-IDF(w, t) is the product of tf(w, t) the number of occurrences of w in t, and a weighting term idf(w) = log [(1 + n) / (1 + df(w))] + 1 where n is the number of text samples in the whole dataset, and df(w) is the number of samples in the dataset that contains w. This weighting procedure dampens the impact of words that occur very frequently in a corpus which may be considered as less informative than those that occur in a small fraction of the corpus. In this work, TF-IDF transforms produced 379-dimensions vectors that were used to train and test classifiers as shown in Figure 1.B.

#### 3.2.2 Transformer models

Each segment of the corpus was first transformed into sequences of word vectors (tokenization) and then to a 768-dimensions vector, using the attentional model CamemBERT, derived from RoBERTa [18], which was trained on a corpus of French texts [17]. CamemBERT was used without fine-tuning. The teenagers’ lexicon was not modified beyond some typos corrections, in order to stay as close as possible to free text writing without complex preprocessing, and to evaluate attentional models robustness when taking into account this population’s specific style (this approach was also used in [12]). It is interesting to note that transformers’ architectures *per se* do not take into account word orders. But, this crucial information is taken into account by combining each word-embeddings with a corresponding positional-embedding. The output of such model is also a 768-dimensions numeric vector that may be fed to any classical classification model or into a perceptron (see Figure 1.A).

### 3.3 Label prediction

Prediction of labels were based on two different methods : the use of classical families of classifiers fed either by TF-IDF or by BERT’s output vectors as described in section 3.2.

The evaluation procedure was the same for each prediction strategy including the use of cross-validation with the same random seed to ensure training and testing dataset comparability, and the use the same performance metrics. For each label, the performance of each prediction strategy was measured by three classical performance metrics in classification. These metrics are respectively : 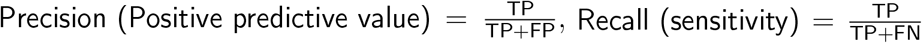 and 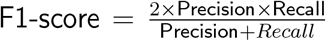, where `TP` are the true positives, `FP` the false positives and `FN` the false negatives counts. The metric `Precision` tells us what proportion of the positive predictions are actually positive. The metric `Recall` tells us what proportion of real positives is correctly classified while the metric `F1-score` corresponds to a measure of balance between the two previous metrics. If either the metrics `Precision` or `Recall` are low, the `F1-score` is low.

#### 3.3.1 Classical models

Several families of classification models were compared, as listed in table 1. Knowing that each family of model is based on their own sets of hyper-parameters, the best model from each one were selected thanks to a 5-folds cross validation procedure on the training dataset with features obtained either by TF-IDF (section 3.2.1) or by attention models (section 3.2.2). The different hyper-parameters of each family of models are listed in table 1. Each pair of models optimized by the cross-validation procedure for each family was finally evaluated on the test dataset. The hyper-parameters optimization and the final model fit on the training dataset was done using `scikit-learn` library [19] excepting gradient boosting classifier which uses `lightgbm` library. Figure 1.A and B illustrate these computations.

**Table 1.** Different families of classical classification methods that are compared with the corresponding list of the hyperparameters. These parameters were optimized by cross-validation for each model family. The grids of values that were tested are available in the following notebooks https://github.com/masedki/ados_familles.

| Classical model families | hyper-parameters |
| --- | --- |
| Elasticnet logistic regression | $C$ and $l_1$ ratio |
| Gradient boosting classifier | Learning rate and number of estimators |
| Random Forest | Maximum of features and bootstrap |
| Support vector classifier | $C$, gamma and kernel |

#### 3.3.2 Transformer model fine-tuning

The comparison of the set of classifiers listed in section 3.3.1 follows two steps. The first step consists in transforming a text segment into a feature vector using the procedures described in sections 3.2.1 and 3.2.2 and a step of choosing and training the model to predict the labels vector from the feature vector obtained in the previous step. In this section, we focus on a model for predicting labels from text in a single step. This was possible by extending `CamemBERT` with a classification perceptron placed at its’ output.

On the top of `CamemBERT` model, a 3-layers perceptron was added in order to predict the 11 labels. An `encoding layer` actually encompasses several neural layers including `self-attention` and *feed-forward* networks. The last layer of the transformer corresponding to the `CLS token` was composed of 768 units, that were progressively reduced to 200, 110 and 11 units, using three perceptron layers placed at the *head* of the transformer. Nonlinear `Tanh` activation function was used for the first two layers (i.e. 768 units to 200, and 200 units to 110), and, for multiple labeling, the last layer (i.e. 110 units to 11). Figure 3 schematizes this architecture.

**Figure 3.**
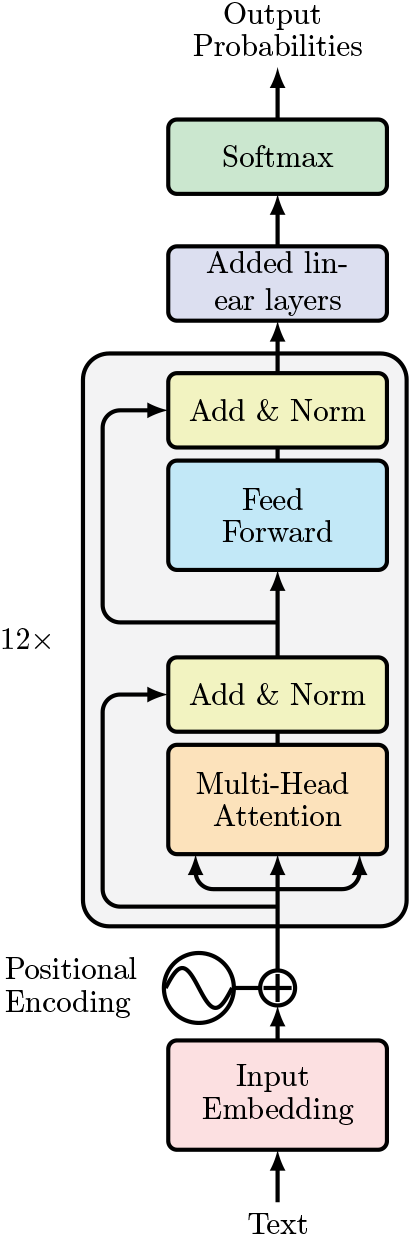
This network summarizes from bottom to top the transformer based neural architecture involved in the section 3.3.2. The block corresponding to CamemBERT is repeated 12 times while the classification network has been added by us in order to carry out the multi-label classification task. The set of weights of the added layers as well as the weights of the last three layers of multi-headed attention at the top of the CamemBERT block are learned on the training set. Indeed, `tune depth` involving three transformer layer blocks was determined by a repeated cross-validation procedure.

A binary cross-entropy loss function (`torch.nn.BCEWithLogitsLoss()`) was used for training in this multi-label-multi-output situation where more than one labels may be found in a single text segment. The weights and the biases of 3-layers perceptron were always back-propagated. The number of transformer’s `encoding layers` that were fine-tuned corresponds to `tune depth` hyper-parameter which was selected using a 5-folds cross-validation procedure which was globally repeated ten times in order to reduce possible variability in the results. The remaining embedding layers below the `tune depth` layer were frozen during the training procedure. The procedure allowing to determine the optimal `tune depth` is schematized in Figure 2.A, and testing of this model with metrics similar to the one used to assess classical classifiers is illustrated in Figure 2.B.

## 4 Results

### 4.1 Fine tuning

Figure 4 represents curve of 5-fold cross validation error repeated ten times as a function of the hyper-parameter `tune depth`. The repetition of the cross-validation procedure was used to remove random effects that would occur during the 5-fold cross validations. Other hyper-parameters were also fixed as described in the notebooks https://github.com/masedki/ados_familles. The main result from this procedure was that best performances were found when the transformer’s parameters were updated from the ninth layer to the perceptron’s output. It is also worth noting that only training the perceptron on top of the transformer penalizes significantly the error rate. Please note that optimization of `tune depth` required one week of processing with a `NVIDIA A100 Tensor Core 40GB GPU`.

**Figure 4.**
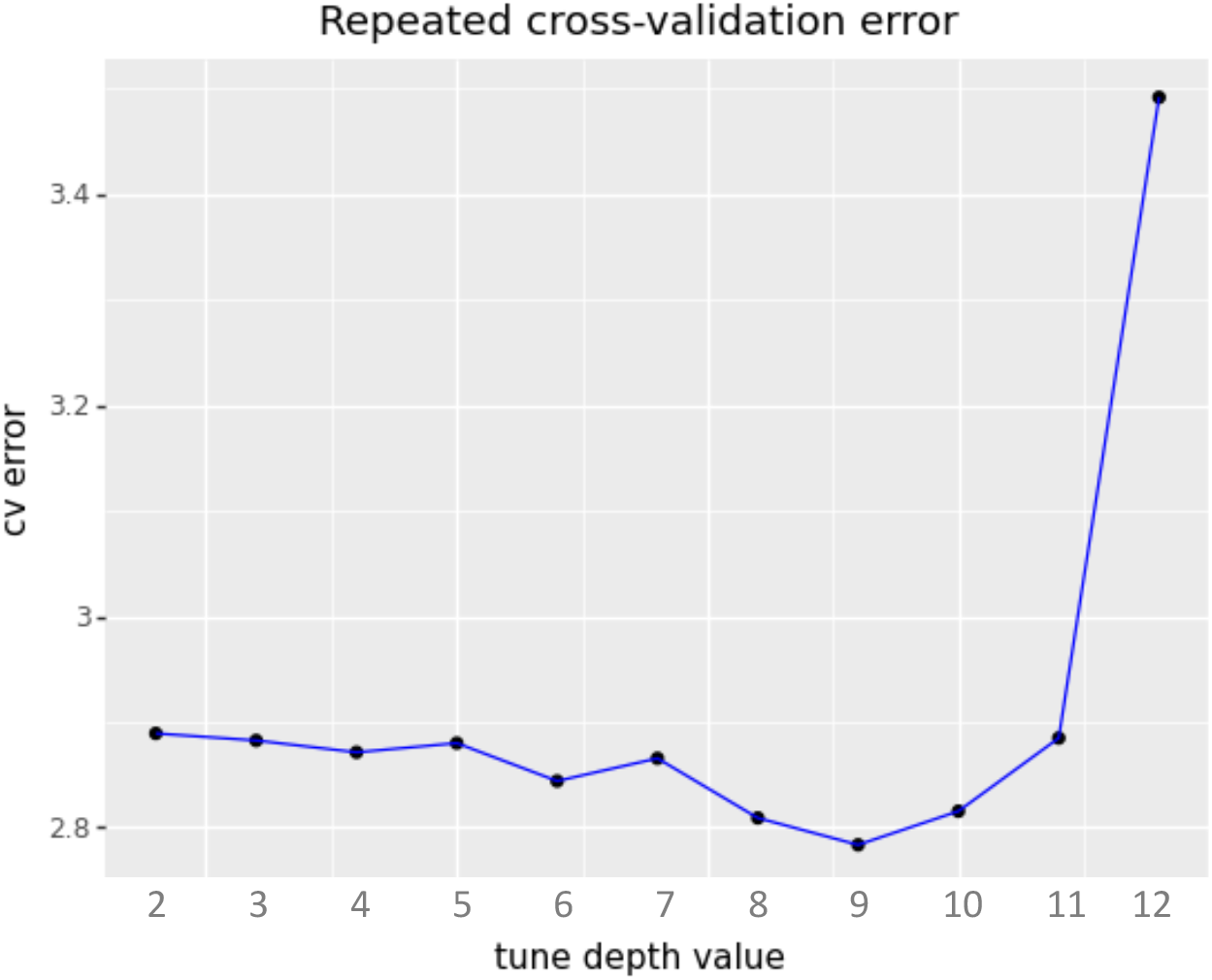
Cross-validation error rate (binary cross-entropy error) as a function of the hyper-parameter `tune depth` during fine tuning. Horizontal axis corresponds to the lower layer to which layer parameters are back-progagated from the output. Twelve corresponds to fine tuning the perceptron only while BERT’s parameters are frozen.

### 4.2 Classification performances

Table 2 summarizes all the results of the numerical computations presented in section 3.3. Overall, the strategy consisting in fine tuning CamemBERT provides the best, yet far from perfect, performances in the majority of the labels. However, concerning valence labels, the best classification performances of fine tuned transformers are found for positive labels in comparison with classical classifiers trained either on BERT output vectors or TF-IDF vectors. Obviously, labels that convey people identity are correctly classified with metrics most of the time superior to 0.9.

**Table 2.** Comparison of different types of error on the test dataset. *lr* stands for *logistic regression* and *SVC* for *Support vector classifier*. The performance of the one-step learning technique described in the section 3.3.2 (Figure 2.B) corresponds to the row labeled `Fine-tuning`. The lines entitled `Elasticnet lr, Gradient Boosting, Random Forest` and `SVC` correspond to the learning methods that we have labeled as classical and described in section 3.3.1. The metrics given in brackets correspond to the TF-IDF text vectorization scenario described in paragraph 3.2.1 (Figure 1.B) while the metrics given outside the brackets correspond to the text vectoring scenario by attention model described in the paragraph 3.2.2 (Figure 1.A). BERT embeddings’ results are written in plain text, tests of classification performances using the same algorithms on TF-IDF sentence vectorization are reported between parentheses. +, -, 0, i : positive, negative, neutral or informative sentences. The persons described in the sentence are labelled the following way. *j* for I, *f* for brother, *s* for sister, *p* for father, *m* for mother, *a* for family member, *t* for other persons

| Label | + | - | 0 | Info | Me | Brother | Sister | Father | Mother | Family | Others |
| --- | --- | --- | --- | --- | --- | --- | --- | --- | --- | --- | --- |
| Support | 72 | 63 | 47 | 146 | 263 | 36 | 57 | 124 | 123 | 72 | 23 |
| Precision |  |  |  |  |  |  |  |  |  |  |  |
| Fine-tuning | 0.67 | **0.85** | 0.40 | **0.78** | **0.86** | **0.91** | 0.97 | **0.97** | **0.94** | **0.91** | 0.78 |
| Elasticnet lr | 0.65(0.69) | 0.71(0.75) | 0.28(0.33) | 0.74(0.61) | 0.85(0.81) | 0.74(0.79) | 0.89(**0.98**) | 0.90(0.93) | 0.83(0.87) | 0.77(0.81) | 0.67(0.75) |
| Gradient Boosting | 0.73(0.65) | 0.88(0.44) | 0.25(**0.45**) | 0.75(0.64) | 0.84(0.80) | 0.75(0.78) | 0.88(0.92) | 0.84(0.93) | 0.79(0.85) | 0.83(0.73) | 0(**0.83**) |
| Random Forest | **0.82**(0.70) | 0.71(0.73) | 0(0.33) | 0.71(0.64) | 0.82(0.80) | 0(0.72) | 0.89(0.92) | 0.86(0.93) | 0.76(0.89) | 0.73(0.77) | 0(0.67) |
| SVC | 0.68(0.70) | 0.79(1) | 0(0) | 0.76(0.66) | 0.85(0.81) | 0.68(0.79) | 0.91(0.94) | 0.90(0.92) | 0.83(0.89) | 0.82(0.79) | 0.50(0.79) |
| Recall |  |  |  |  |  |  |  |  |  |  |  |
| Fine-tuning | **0.82** | 0.37 | **0.38** | **0.81** | 0.97 | **0.89** | **0.98** | **0.93** | **0.93** | **0.86** | **0.61** |
| Elasticnet lr | 0.56(0.49) | **0.40**(0.10) | 0.11(0.02) | 0.73(0.75) | 0.94(1) | 0.39(0.64) | 0.68(0.81) | 0.76(0.85) | 0.82(0.85) | 0.67(0.64) | 0.26(0.52) |
| Gradient Boosting | 0.49(0.49) | 0.22(0.11) | 0.02(0.11) | 0.75(0.66) | 0.97(0.95) | 0.17(0.81) | 0.49(0.86) | 0.70(0.85) | 0.69(0.90) | 0.47(0.64) | 0.00(0.43) |
| Random Forest | 0.38(0.43) | 0.08(0.17) | 0(0.04) | 0.75(0.59) | 0.96(0.92) | 0.00(0.86) | 0.30(0.95) | 0.52(0.92) | 0.60(0.93) | 0.33(0.71) | 0.00(0.52) |
| SVC | 0.57(0.46) | 0.30(0.05) | 0(0) | 0.77(0.73) | 0.94(**1**) | 0.42(0.64) | 0.72(0.86) | 0.77(0.78) | 0.81(0.89) | 0.64(0.58) | 0.09(0.48) |
| F1-score |  |  |  |  |  |  |  |  |  |  |  |
| Fine-tuning | **0.74** | **0.51** | **0.39** | **0.79** | **0.91** | **0.90** | **0.97** | **0.95** | **0.93** | **0.89** | **0.68** |
| Elasticnet lr | 0.60(0.57) | 0.51(0.17) | 0.15(0.04) | 0.74(0.67) | 0.89(0.89) | 0.51(0.71) | 0.77(0.88) | 0.82(0.89) | 0.83(0.86) | 0.72(0.71) | 0.38(0.62) |
| Gradient Boosting | 0.58(0.56) | 0.35(0.18) | 0.04(0.17) | 0.75(0.65) | 0.90(0.87) | 0.27(0.79) | 0.63(0.89) | 0.77(0.89) | 0.74(0.88) | 0.60(0.68) | 0(0.57) |
| Random Forest | 0.51(0.53) | 0.14(0.28) | 0(0.08) | 0.73(0.61) | 0.89(0.86) | 0.00(0.78) | 0.45(0.93) | 0.65(0.92) | 0.67(0.91) | 0.46(0.74) | 0(0.59) |
| SVC | 0.62(0.55) | 0.44(0.09) | 0(0) | 0.77(0.69) | 0.90(0.89) | 0.52(0.71) | 0.80(0.90) | 0.83(0.84) | 0.82(0.89) | 0.72(0.67) | 0.15(0.59) |

## 5 Discussion

In the present work, we aimed at testing the feasibility of using automatic language processing methods based on transformers to categorize the writings of French adolescents on their family relationships. Based on previous research, we considered that relational valence could constitute a relevant element as a psychological outcome. We used transformers because these models are pre-trained on large corpora allowing us to benefit from the “general” linguistic and semantic knowledge encoded inside and to fine-tune them on smaller datasets of labeled sentences. We wanted to see if a model recently made available in French would have sufficient semantic representation capacity to determine valence, as in sentiment analysis, as well as to identify the people described in the texts. To begin with the technical aspects associated with learning the 11 labels, the hyper-parameters selected are consistent with published studies employing BERT or its variants.

In this study, we find that models based on fine-tuning the inner layers of the transformer outperform those based on classifying the output vectors of the transformer head. Other works have also reported that intervening in the internal structure of BERT could have a benefit, although this strategy is debatable for that it separates the new fine-tuned model from the original one. The question was whether there is a level of depth to which the backpropagation of the error must access to maximize performance. A Study on BioBERT model to classify multiple clinical concepts has shown that freezing up to six bottom layers of the encoder during training maintained good performances [20]. In the present work, best error rate over validation set could be found around the ninth layer. It can be suggested that the learning depth corresponding to better prediction performances informs us about the type of information that are processed along the different layers of the transformer. Van Aken et al. raise the hypothesis that different types of processing and representation exist within transformer networks and conclude that “it could be beneficial to fit parts of the network to specific tasks in pre-training, instead of using an end-to-end language model task”[16]. Although our approach (selecting the depth of fine-tuning) differs technically from theirs (selecting the output of an internal layer of the transformer), we concur in the idea that these pre-trained models have an internal architecture whose knowledge would help optimizing new tasks.

The predictive performance of the fine-tuned model concerning either the relational semantics of sentences or the identity of persons was compared with that of classical classification algorithms. The first result is that the last layer of BERT conveys information about the identity of individuals at higher performances (i.e. larger F1-scores) by the SVC algorithm compared to Elastic Net, Gradient Boosting and Random Forest. Moreover, BERT fine-tuning, as described above, brings a substantial gain on precision and recall compared to classical algorithms. Fine-tuned BERT is also found better when compared with classical classifiers fed with TF-IDF vectors instead of word-vectors embeddings. This suggests that transformers have a sufficient power of representation of subjects, in terms of categories of people, to deal with relational situations. As such, this result did not solve a real methodological issue and does not impose this technology as efficient to detect these entities because simple parsing algorithms can easily do so. But it shows that transformers have the capability of making it possible to embed the information of the agents being represented in the texts in combination with other semantic information for any purpose and learning.

Now regarding semantic labels representing valence, we obtain contrasted prediction performances. Compared with classical algorithms, the best F1-scores were obtained using BERT fine-tuning for the +, -, 0 and “info” labels (i.e. positive, negative, neutral and information labels). However, only + (positive) and “info” labels were reached 0.7 F1-scores, with a better Recall than Precision measures. As it stands, the proposed fine-tuning method, on a small sample of texts, presents a better recall capacity for positive and informative texts. This approach may be used for research in large corpora of texts and aiming at extracting the maximum number of texts dealing with positive relations or even distinguishing them from informative texts. However, it is interesting to note that the - (negative) label is associated with a better precision. This can also have the advantage, in large text corpora, of targeting selectively negative texts with a reduced number of false positives. In any case, if the precision/recall profiles of the labels turn out to be distinct, it seems necessary to conduct additional investigations to see if their use should be thought in distinct scenarios of use.

### 5.1 Limitations of the study

In this study we used a small learning base and tried to use it to train sophisticated natural language processing models. These models being pre-trained on very large textual databases were to benefit from their ability to represent relevant semantic information. The unfavorable results concerning the recall of the negative valence labels are possibly related to the small size of the training dataset. We conclude that a substantial effort to build larger databases of labels realized by human operators could help progressing. Concerning the labeling of complex psychopathological criteria by experts, this may lead to quite expensive work. If such databases are created, the possibilities of translating them automatically from one language to another or of using multilingual models will have to be evaluated.

## 6 Conclusion

In this paper, we propose the use of recent natural language processing methods in child and adolescent psychiatry. To our knowledge, no study has previously investigated the possibility of labeling texts related to family relationships using these methods. Our work brings contrasted preliminary results notably by showing that labels concerning positive relational valence are predicted with better precision and recall. On the other hand, negative valences are more complicated to label and our results do not provide an immediate solution to detect difficult family situations. Further work should improve the model in order to meet the requirements of clinical use. Nevertheless, the use of these methods, despite their limited predictive power, should be considered for large-scale investigations of internet and social media databases to characterize the evolution of young people’s views of their family relationships. Finally, we concur with Abbe and colleagues, on the idea that new text mining techniques might discover new variables from the clinical experiences reported directly by the patients [21]. Beyond “classical applications” such as diagnosis or suicide prediction, one could propose automatizing of validated clinical measures or psychological constructs. However, to achieve these goals, important efforts to constitute properly labelled text corpus would be necessary.

## Data Availability

Computer code is available online.

https://github.com/masedki/ados_familles

## 7 Acknowledgments

We wish to warmly thank Christine Hassler, Muriel Letrait, Guillaume Macher, François Marty, Elsa Ramos, Anne Revah-Lévy, Philippe Robert, and françois de Singly, for conceiving the multidisciplinary protocol to gather the adolescents’ texts dataset used in this study and published in [2].

## 8 Ethical statements

This study reports an additional analysis of data collected and published in [2] which obtained ethical agreement from the CCTIRS, CNIL (national IRB for this kind of study) with number MG/CP 10962.

